# Effects of Aerobic Exercise on Brain Age and Health in Older Adults: A Single-Arm Clinical Trial

**DOI:** 10.1101/2022.06.13.22276337

**Authors:** An Ouyang, Can Zhang, Noor Adra, Ryan A. Tesh, Haoqi Sun, Dan Lei, Jin Jing, Peng Fan, Luis Paixao, Wolfgang Ganglberger, Logan Briggs, Joel Salinas, Matthew Bevers, Christiane Wrann, Zeina Chemali, Gregory Fricchione, Robert J. Thomas, Jonathan Rosand, Rudolph E. Tanzi, M. Brandon Westover

## Abstract

**Backgrounds:** Exercise is an attractive, widely accessible intervention to promote cardiovascular health; however, evidence that exercise improves brain health is sparse. Here, we hypothesized that aerobic exercise would improve brain health of sedentary older adults, as reflected by cognition health, sleep macro- and micro-architecture, and brain age index (BAI), a biomarker of brain health derived from the overnight sleep electroencephalogram (EEG).

**Methods:** Sedentary older adults were recruited to complete a 12-week aerobic exercise. Home wearable devices were used to monitor heart rate and overnight sleep EEG over the period. NIH Toolbox Cognition Battery, in-lab overnight polysomnography, cardiopulmonary exercise testing and multiplex cytokine assay were employed to determine pre- and post-exercise brain health, exercise capacity and plasma proteins.

**Results:** 26 participants completed the initial assessment and exercise program, and 24 completed all procedures. Participants significantly increased maximal oxygen consumption (VO2max) and decreased resting and sleeping heart rate after the exercise regimen. Cognition performances were significantly improved following the exercise program while no significant differences were seen in BAI and sleep macro- and micro-architecture. Plasma IL-4 was elevated while IL-8 was reduced after the exercise regimen. Home sleep data revealed a 3.59% increase in the percentage of N3 sleep over a 12-week.

**Conclusions:** We conclude that cognitive function and N3 sleep were improved by a 12-week moderate-intensity exercise program in sedentary older adults, associated with improvements in VO_2_max and plasma cytokine profiles. Our data show the value of integrating multi-modal assessments to study the effect of brain health targeted approaches.

**Funding:** Dr. Westover received support during this work from the McCance Center for Brain Health, the Glenn Foundation for Medical Research and the American Federation for Aging Research through a Breakthroughs in Gerontology Grant; through the American Academy of Sleep Medicine through an AASM Foundation Strategic Research Award; by the Football Players Health Study (FPHS) at Harvard University; from the Department of Defense through a subcontract from Moberg ICU Solutions, Inc, and by grants from the NIH (R01NS102190, R01NS102574, R01NS107291, RF1AG064312, R01AG062989, R01AG073410), and NSF (2014431). Dr. Wrann was supported by a SPARC Award from the McCance Center for Brain Health. Dr. Tanzi and Dr. Zhang were supported by the Cure Alzheimer’s Fund.

**Clinical trial number:** National Clinical Trial: NCT04210882

**One Sentence Summary:** We observed that exercise improved slow wave sleep, increased circulating neuroprotective cytokines and improved cognition health in older adults.

## Introduction

Sleep and brain health are tightly linked, with improved sleep generally correlating with objective and subjective measures of improved brain structure and function. Changes in both sleep macro- and micro-architecture are associated with aging and with general health [1]. These aging-associated sleep disturbances are independent risk factors that contribute to both impaired cognitive function [2-5] and mortality in older adults [6, 7]. There is a critical need for 1) non-pharmacological approaches to improve sleep and thus positively impact brain health, and 2) practical and precise ways to track the effect of interventions that aim to improve brain health.

Exercise is an intervention of key interest, as accumulating evidence shows beneficial effects on both sleep quality and on measures of cognitive brain health in older adults [8-10]. Previous work has suggested that exercise increases slow wave sleep (SWS) and fast-sigma power and reduces wake time [11-14]. However, other studies have found minimal effects of exercise on deep sleep [15, 16]. In addition, it remains uncertain whether exercise improves cognitive function and sleep quality in older adults with dementia [17, 18]. Recent AAN (American Academy of Neurology), ACSM (American College of Sports Medicine) and WHO guidelines recommend that elder individuals exercise 150-300 minutes weekly to maintain brain health [19, 20]. However, the optimal frequency, intensity, time, and type (FITT) of aerobic exercise are debatable. Overall, the differences in exercise regimen design, target population, and sleep measurements in these previous exercise intervention trials leave open the question of whether and to what extent exercise is able to improve brain health.

Brain age is a construct based on an objective measure of brain structure or function, such as magnetic resonance imaging [21-25] or sleep electroencephalography [26-28]. The Brain Age Index (BAI) is the estimated “brain age” minus chronological age and reflects biological age (“older” or “younger” than expected for chronological age). A previously proposed sleep-EEG based BAI [26] is increased in individuals with neuropsychiatric or cardiometabolic diseases [29] and in individuals with mild cognitive impairment (MCI) or dementia [30] and associated with increased mortality [31]. However, no study has evaluated whether sleep EEG-based BAI can be lowered through interventions that improve brain health.

In this study, our primary goal was to investigate whether an exercise regimen designed to improve aerobic fitness could improve brain health as reflected by improvement in cognition, reduction in BAI, and enhancement of key features of sleep macro- and micro-architecture. A secondary goal was to assess changes in plasma cytokines as plausible mechanisms mediating exercise effects on sleep and cognition. We therefore conducted an interventional study of sedentary older adults testing two hypotheses:1) a 12-week, 150 minutes-weekly moderate-intensity aerobic exercise program will improve brain health, measured by performance on cognitive assessments, sleep EEG based-BAI, and specific improvements in sleep macro- and micro-architecture; 2) improvements in BAI and cognitive performance will correlate with improvements in aerobic fitness, defined by changes in maximal oxygen consumption (VO_2_max) and plasma cytokine biomarkers.

## Methods

### Study design and participants

We conducted a before–after (pre–post) interventional study. Adults aged 50 to 75 years old were recruited from the greater Boston area through online advertisements, flyers, and outpatient clinics at the Massachusetts General Hospital. Participants were included if sedentary (average time spent exercising was less than 60 minutes per week in the past 6-months) and received clearance by their primary care or personal physician to participate in the 12-week moderate-intensity exercise program. A pre-screening survey was used to select eligible participants with the following exclusion criteria: 1) history of neurological illness (e.g. poorly controlled epilepsy with >1 seizure per month in the last 6-months, stroke with residual motor language deficits, multiple sclerosis, Parkinson’s disease, dementia, head trauma in the past 6-months with loss of consciousness >30 min, cerebral palsy, brain tumor, normal-pressure hydrocephalus, or Huntington’s disease); 2) history of untreated or poorly controlled neuropsychiatric illness; 3) diagnosed with moderate or severe sleep apnea (apnea-hypopnea index ≥ 15/hour of sleep) or using a continuous positive airway pressure machine (CPAP); 4) HIV infection; 5) history of falling within the past 6-months; 6) inability to safely exercise or perform any of the tests, e.g. failure to complete: cardiopulmonary exercise testing (CPET) testing (or development of cardiac symptoms during CPET), or lack of English proficiency which limited cognitive testing.

Power analysis suggested that a minimum of 34 subjects (80% power, 0.5 mean of paired differences, 1.0 standard deviation of differences [32]) would be needed to evaluate whether a 12-week moderate intensity exercise program can improve BAI and cognitive scores. We proposed to recruit 35 subjects to complete all assessments over 1 year period to establish adequate power and to minimize Type II error.

Written informed consent was obtained prior to participation in the study, which was approved by the Mass General Brigham Review Board (National Clinical Trial: NCT04210882). On the day of consent, participants completed CPET, cognitive testing, and a blood draw. Participants returned to the hospital within one week to undergo a diagnostic polysomnography (PSG). These procedures were repeated at the end of the study. After the PSG, participants were instructed to begin a program of moderate-intensity exercise. Details of the exercise program are described below. Study design is shown in Figure 1A.

**Figure 1.**
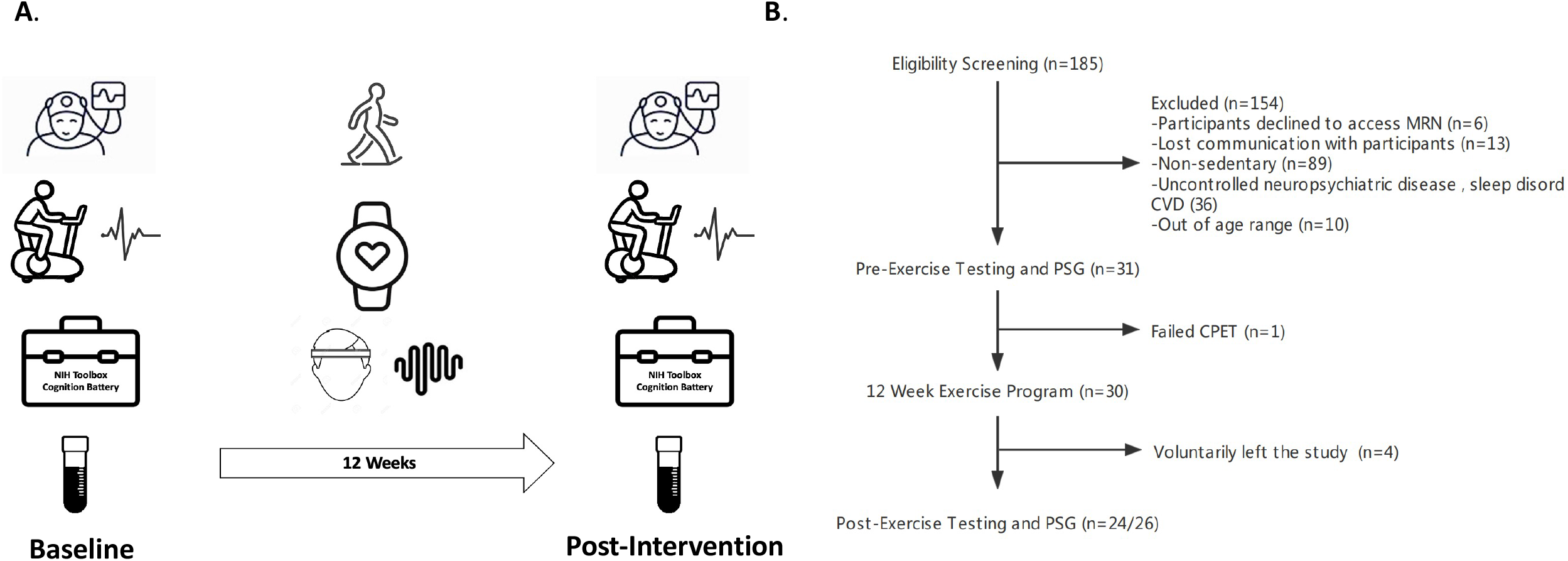
Study design (Fig. 1A) and enrollment flow chart (Fig. 1B). In Fig. 1A, left/right column pictures from top to bottom are polysomnography (PSG), cardiopulmonary exercise testing (CPET), NIH toolbox cognition battery and blood samples. The middle column pictures from top to bottom are exercise, physical activity tracker and home sleep headband.

### Cardiopulmonary Exercise Testing (CPET)

Aerobic exercise capacity, resting heart rate and cardiac-event risk were measured at the Brigham and Women’s Hospital CPET laboratory by the clinical exercise physiologist. Prior to the visit, participants were instructed to refrain from smoking, eating, and drinking caffeinated or alcoholic beverages for an 8-12h period overnight. During the exam, participants’ VO_2_max was determined by the following ramp cycle protocol: cycle ergometer (Lode Corival, Netherlands) intensity was set to 10-25 watts/minute (depending on the subject’s capacity) at the speed of 60 rpms until exhaustion or clinical concerns were voiced. Respiratory measurements and electrocardiogram (ECG) were recorded by the Ultima CPX system (Medgraphics, MN). For two participants who refused to complete the final assessment of their VO_2_max due to COVID-19 concerns, VO_2_max was determined by a 1-mile walking test in an outdoor track. Briefly, participants were instructed to walk as quickly as possible while avoiding jogging or running. Post walking heart rate, walk time, body weight, age, and sex were used to calculate the VO_2_max as described before [33].

### Cognitive Testing

The National Institute of Health (NIH) Toolbox Cognition Battery was administered in a quiet and private environment by a trained study coordinator [34]. This battery is one of the core domains of the NIH Toolbox for Assessment of Neurological and Behavioral Function and consists of seven instruments. Fluid cognition is a measure of five of these instruments (Dimensional Change Card Sort (DCCS), Flanker Inhibitory Control & Attention (FICA), List Sorting Working Memory (LSWM), Pattern Comparison Processing Speed (PCPS), and Picture Sequence Memory (PSM) whereas crystallized cognition is a composite of two of these instruments (Picture Vocabulary Test (PVT) and Oral Reading Recognition (ORR). Total cognition is a composite of fluid and crystallized scores. Absolute for each of the seven tests and the three composite scores were used for analyses (all non-age adjusted).

### Plasma biomarkers level assessment

The morning of the CPET appointment, overnight fasting blood samples were drawn by a phlebotomist at the Mass General Brigham Center for Clinical Investigation. Blood was centrifuged at 2000g for 15 minutes. Plasma was aliquoted and stored in a -80° freezer for protein analysis. The levels of cytokines were measured using the established electrochemi-luminescence-based multi-array method through the Quickplex SQ 120 system (by the Meso Scale Diagnostics LLC) using previously reported methods [35-37]. In brief, the system utilizes a 96-well-based high-throughput readout. The human proinflammatory multi-plex kits were utilized which enabled detection of 8 cytokines in our samples, including INF-γ, TNF-α, IL-2, IL-4, IL-6, IL-8, IL-10, and IL-13. Procedures for measuring cytokines levels followed the manufacturer protocols. In brief, our samples and standard proteins provided by the manufacturer were prepared and incubated at 4°C overnight. Then the mixed solutions were placed on a shaker for 2 hours, followed by washing and then incubation with detection antibodies for another 2 hours. Next, the electrochemi-luminescence signals were captured through the SQ 120 system. Finally, protein concentrations (pg/mL) in the samples were calculated using the manufacturer-provided standard concentrations.

### Diagnostic polysomnography (PSG)

PSG was performed at the American Academy of Sleep Medicine (AASM)-Accredited Massachusetts General Hospital Sleep Disorders Unit. During the COVID pandemic, all participants were cleared with PCR COVID testing 72 hours prior to the study. A minimum of six hours of overnight sleep was monitored using conventional in-lab polysomnography (Compumedics, NC) with 250 Hz sampling rate and a 0.3-35 Hz bandpass filter. EEG data was recorded from frontal (F3 and F4), central (C3 and C4), and occipital (O1 and O2) electrodes, and then referenced to contralateral mastoid electrodes (M2 and M1). Electrooculogram (EOG), electromyogram (EMG), ECG, pulse oximetry, respiration, and nasal flowmeter were used to assess sleep apnea and leg movements. Sleep was staged as non-rapid eye movement (NREM) stages 1 to 3, R (REM), or awake (W) stages in consecutive 30 second epochs following AASM criteria by Registered Polysomnographic Technologists [38]. Two participants who completed the initial assessment and exercise program declined to complete the final PSG assessment because of COVID concerns. Their final sleep EEGs were recorded by a home device.

### 12-week exercise regimen

Participants completed a 12-week exercise regimen designed as follows: maintenance of 50-75% maximal heart rate (HRmax = 220 - age) intensity for 30 minutes per day, 5 days per week. Participants began with 15 minutes/day x 3 days in the week 1, 20 minutes/day x 4 days in the week 2, then 30 minutes / day x 5 days over weeks 3-12. If unable to exercise for 30 minutes continuously, participants were allowed to split the 30 minutes of exercise into sessions of 10-15 minutes with 5-minute rest intervals. For feasibility, the type and timing of exercise were not restricted, however participants were encouraged to exercise around the same time each day. Participants were given a physical activity tracker (Fitbit, USA) to monitor their exercising heart rate to ensure the exercise time and intensity met the intensity criteria. Additionally, participants were instructed to log every physical activity session in case of device issue. The study team met in person or via teleconference (during the COVID-19 pandemic) with participants bi-weekly to answer questions.

### Home data collection

Participants were required to wear the Fitbit 24×7 at their maximal convenience and sync the device every 5 days to upload the data to the cloud, allowing the study team to check adherence to the exercise program. Additionally, overnight sleep EEG was recorded using a home sleep headband (Dreem 2, France; or Prodigy, Canada) two nights per week throughout the 12-week exercise program. The Dreem 2 headband records via five EEG dry electrodes (Fpz, F7, F8, O1, and O2) at 250 Hz sampling rate with a 0.4–35 Hz bandpass filter. Sleep staging of Dreem 2 headband was performed using an automated algorithm [39]. The Prodigy sleep monitor records via six snap forehead electrodes (left/right frontal EEG sensors, left/right EOG sensors, mastoid and chin sensors) at 120 Hz sampling rate with 0.33-35 Hz bandpass filter [40]. Sleep staging of Prodigy headband was performed using a previously described automated algorithm [41]. Heart rate and EEG data were examined by the study team weekly to ensure device adherence.

### Exercising time and sleeping heart rate

Continuous heart rate was tracked using a Fitbit, which measures heart rate via an optical sensor every 10 to 15 seconds. We set three timepoints to determine the time of exercise: T1 (peak heart rate timepoint), T2 (45 minutes before T1), and T3 (45 minutes after T1). Periods of exercise sessions were defined as compliant with the study protocol when the heart rate during T2 and T3 was ≥50-75% HRmax for at least 10 minutes. The sum of all such sessions was defined as the total exercise time. In the absence of Fitbit data, the exercise log was then used. Fitbit automatically records sleep and wake timepoints. Sleeping heart rate was defined as the average heart rate during the sleeping period.

### EEG preprocessing and artifact removal

For both in-lab PSG and at-home sleep measurements, EEG signals were notch-filtered at 60Hz to reduce line noise and bandpass filtered from 0.5Hz to 20Hz to reduce myogenic artifacts. The signals were then segmented into 30-second epochs. Epochs with artifacts were excluded in two steps. In the first step, we removed “definite” artifacts by excluding epochs with a maximum absolute amplitude larger than 500µV or a flat (standard deviation < 1µV) amplitude that lasted 2 seconds or longer. In the second step, for the remaining epochs, we trained a linear discriminant analysis (LDA) classifier to classify each epoch into artifact vs. not artifact. We used the total power and 2^nd^ order difference (for abrupt non-physiological changes. see supplemental material) of the spectrum as inputs to the LDA classifier. To train the classifier, we manually labeled each epoch in randomly selected EEGs (two channels: F7-O1, F8-O2) with different ratios of definite artifact per epoch across the night: 10 EEGs with a ratio of 25-50%; 10 EEGs with a ratio >50%. After training, the LDA classifier labeled each epoch as artifact or no artifact. By comparing these with manually assigned labels, we derive a receiver operating characteristic (ROC) curve (Fig. S1). We used this ROC curve to find a threshold that achieved a false negative rate (FNR) of 10%, as visual analysis of this cutoff produced an acceptable tradeoff between retaining high quality signal and rejecting artifactual epochs.

### Brain age computation and spindle analysis

The BAI was computed as described previously [29]. Briefly, BAI was calculated using a machine learning model with overnight sleep EEG features. For each 30-second epoch, we extracted 96 features from both time and frequency domains, including line length (signal complexity), signal kurtosis (extent of extreme values); max, min, mean, and standard deviation of relative delta, theta, alpha band powers, and delta-to-theta, delta-to-alpha, and theta-to-alpha power ratio computed across 2-second sub-epochs; and kurtosis of spectral power in delta, theta, alpha, and sigma bands. We averaged these features within each of the five sleep stages over time, concatenated them, to finally arrive at 96×5 = 480 features to represent physiologic information in a night of sleep. Sleep EEG data with an artifact ratio < 0.5 was computed for the BAI. In addition, sleep spindles, delta or slow oscillation (0.2-4.5Hz), and their coupling during N2 stage were detected and their summary features were quantified using the Luna software[42]. Spindle features included spindle density, average peak frequency, average amplitude, and average duration. The delta or slow oscillation features include their density, average amplitude, average duration, positive-to-negative slope, and negative-to-positive peak. The coupling features included magnitude of coupling, number of spindles overlapping a detected delta or slow oscillation, and average delta or slow oscillation phase at the spindle peak.

### Statistical analysis

Baseline and post-exercise metrics, including BAI, sleep macro- and micro-architecture, cognitive scores, VO_2_max, and heart rate, were analyzed with paired t-tests. Effect size (ES, Cohen’s d) was calculated as mean change of pre-exercise and post-exercise divided by standard deviation of pre-exercise [43]. Associations were measured using Pearson’s correlation coefficients [44]. Robust linear regression (Huber) [45] was used to evaluate trends in the 12-week data recorded by the home devices. A linear mixed effects model was employed to examine the effects of age, gender, BAI, VO_2_max, heart rate, cognitive scores, and cytokines. Data is presented as mean ± standard error (SE), except otherwise specified. Statistical significance was defined as p<0.05. Statistical analyses were performed with Python 3.7 (Python Software Foundation), Rstudio (RStudio Team 2020), and GraphPad Prism 8.0 (San Diego, CA).

## Results

### Participant characteristics

Thirty-one eligible participants were enrolled between November 2019 and June 2021 (Figure 1B). Five participants were subsequently excluded: one was withdrawn because of cardiovascular risk detected by CPET; two were withdrawn for personal reasons related to the COVID-19 pandemic; 2 were due to insufficient access to exercise resources during the pandemic. Of the remaining twenty-six participants, two declined to complete the final in-lab PSG and CPET assessments due to COVID concerns. Their final sleep EEG and VO_2_max were assessed outside of the hospital (see above *Methods*). Prior to reaching the pre-defined target sample size of 34, recruitment was discontinued due to unanticipated enrollment and financial challenges of the COVID-19 pandemic. Thus, all analyses reported herein are based on the participants who completed the study (N = 26).

Our final sample included 20 females and 6 males with an average age of 60 ± 7.37 years (Mean ± Standard Deviation, SD). All subjects completed at least college level education, averaging 17.19 ± 2.8 (Mean ± SD) years of education. The average body mass index (BMI) was not significantly different between pre- and post-exercise time points. Average apnea-hypopnea indices were 4.05 ± 3.92 (Mean ± SD) and 3.06 ± 3.94 (Mean ± SD) at initial and final assessments. Participants exercised for approximately 47 ± 13 days (Mean ± SD) over the 12-week period and 55 ± 9.8 minutes/day (Mean ± SD), based on their heart rate data and exercise logs. Participant characteristics are presented in Table 1.

**Table 1.** Characteristics of Participants. Participant Characteristics. YOE: years of education. BMI: body mass index. AHI: apnea–hypopnea index. Exercise time and days were counted when heart rate was 50%-75% of max heart rate (220-Age). One participant selected more than one race. Data is mean ± standard deviation.

|  | Pre-Exercise | Post-Exercise |
| --- | --- | --- |
| Age | 60.11 ± 7.37 | - |
| Sex (F, M) | 26 (20, 6) | - |
| Race | 1 (American India or native Alaska)<br>3 (Asian)<br>1 (Black or African)<br>23 (White) | - |
| YOE(SD) | 17.19 ± 2.80 | - |
| BMI (SD) | 25.86 ± 4.16 | 25.83 ± 3.97 |
| AHI | 4.05 ± 3.92 | 3.06 ± 3.94 |
| Exercise days | - | 47 ± 13 |
| Exercise time (minutes) | - | 55 ± 9.8 |

### Physical fitness

As expected, the 12-week aerobic exercise program improved measures of cardiovascular fitness, as measured by VO_2_max (Pre-Ex: 21.11 ± 1.03 ml/kg/min, Post-Ex: 22.39 ± 1.12 ml/kg/min; p= 0.0413; Figure 2A). Relative to baseline measurements, decreases in resting heart rate (Pre-Ex: 66.66 ± 0.83 bpm, Post-Ex: 65.13 ± 0.61 bpm, p= 0.011; Figure 2B) and sleeping heart rate (Pre-Ex: 64.55 ± 1.28 bpm, Post-Ex: 62.93 ± 1.03 bpm; p=0.028; Figure 2B) were also observed following the exercise program.

**Figure 2.**
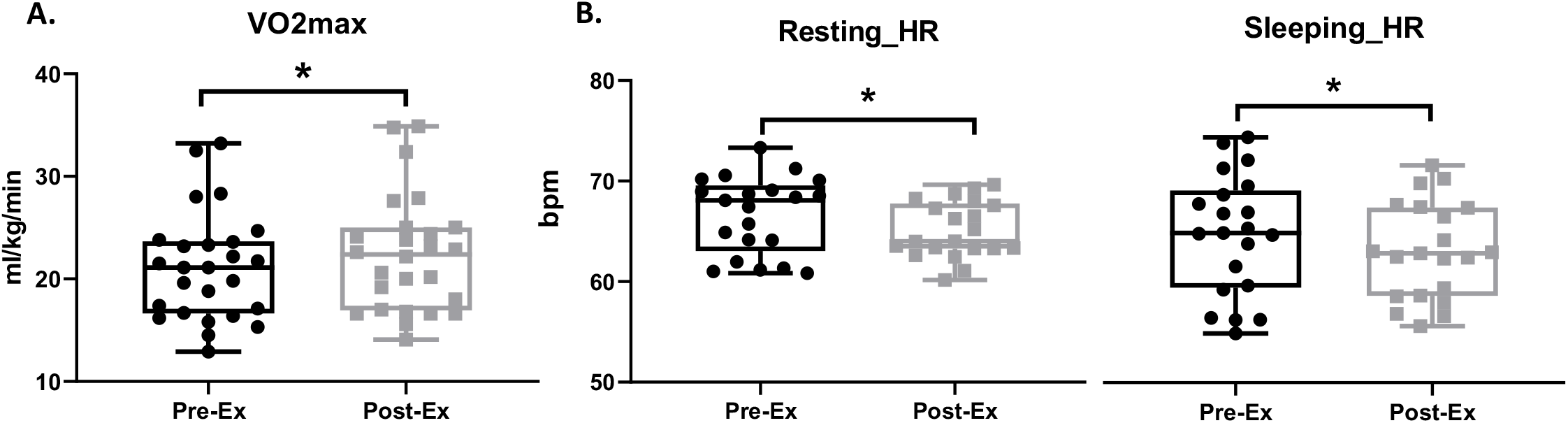
Comparison of baseline (pre-ex) and after 12-weeks of exercise (post-ex) cardio fitness data via measurement of maximum oxygen consumption (VO_2_max, Fig. 2A), resting heart rate (Resting HR) and sleeping HR (Fig. 2B). N=26, data is mean ± standard Error. * is set as statistically significant, p<0.05.

### Cognition performance score

After the 12-week exercise program, brain cognitive performance was improved in multiple domains, including the composite domains of crystallized intelligence (Pre-Ex: 115.1 ± 1.4, Post-Ex: 117.4 ± 1.38; ES= 0.3, p= 0.0008; Figure 3A), fluid intelligence (Pre-Ex: 104.8 ± 2.35, Post-Ex: 109.3 ± 2.23; ES= 0.38, p= 0.0058; Figure 3B) and total cognition (Pre-Ex: 111.1 ± 1.70, Post-Ex: 115.2 ± 1.60; ES= 0.48, p= 0.0001; Figure 3C). Additionally, improvements in the subdomains of processing speed, as measured by Pattern Comparison Processing Speed (Pre-Ex: 100.7 ± 3.92, Post-Ex: 107.6 ± 3.52; ES= 0.36, p= 0.036; Figure. 3D), and language skills, as measured by Oral Reading Recognition (Pre-Ex: 113 ± 1.19, Post-Ex: 116.5 ± 1.13; ES= 0.59, p<0.0001; Figure 3E), were observed. Improvement in the List Sort Working Memory Test was marginally significant (Pre-Ex: 104.2 ± 1.56, Post-Ex: 107.8 ± 9.11; ES= 0.42, p=0.0615; Figure 3F).

**Figure 3.**
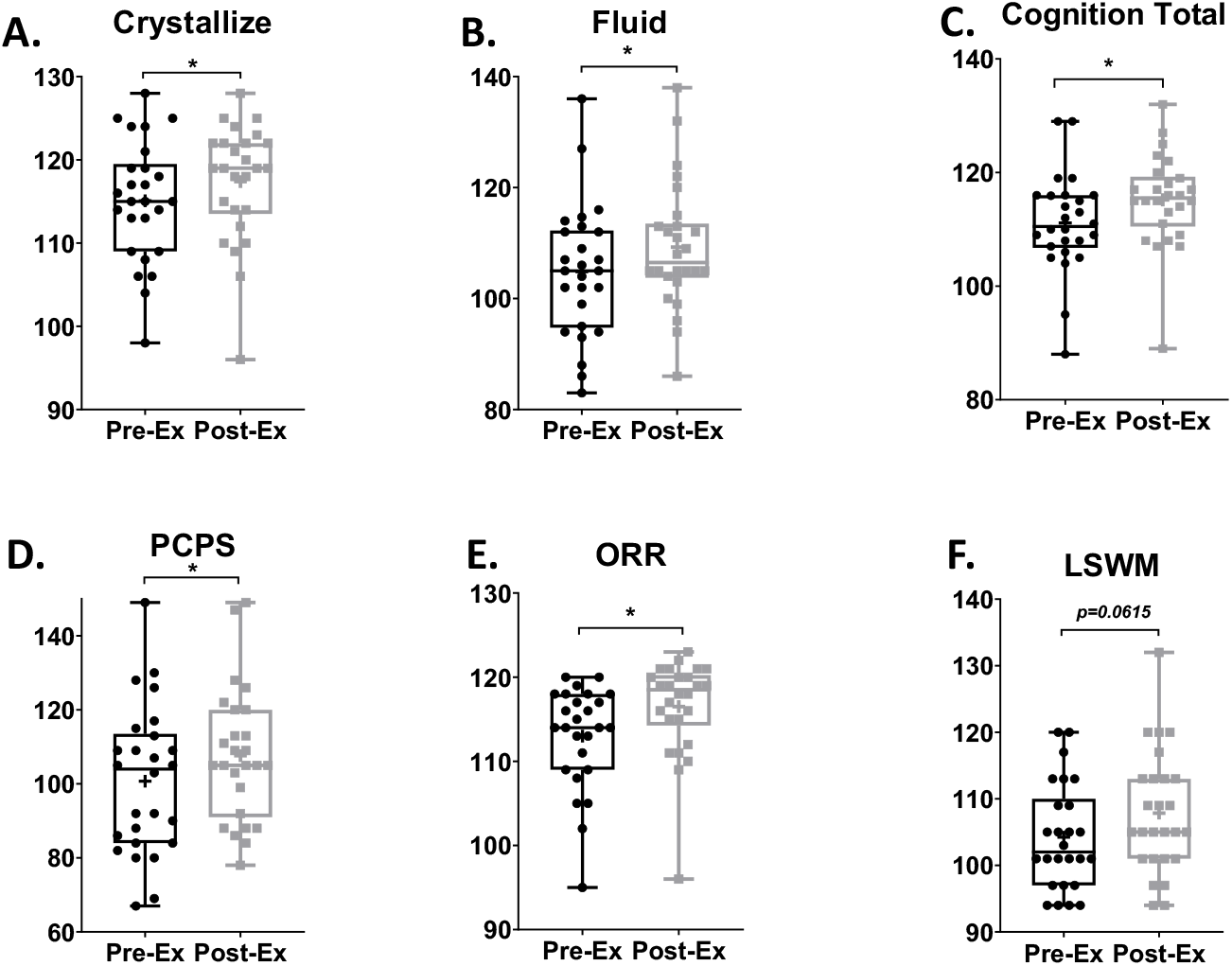
Comparison of baseline (pre-ex) and after 12-weeks of exercise (post-ex) cognitive function scores. Crystallize cognition score (Fig. 3A); Fluid cognition score (Fig. 3B); Total cognition score (Fig. 3C). Pattern comparison processing speed score (PCPS, Fig. 3D); Oral reading recognition score (ORR, Fig. 3E); List sort working memory score (LSWM, Fig. 3F). N=26, data is mean ± standard error. * is set as statistically significant, p<0.05.

### Plasma biomarkers

Compared to the baseline level of plasma cytokines, IL-8 level at post-exercise was significantly decreased (Pre-Ex: 5.50 ± 0.51 pg/ml, Post-Ex: 4.34 ± 0.33 pg/ml; ES= 0.56, p=0.0022, Figure 4A). After 12-week moderate-intensity exercise, plasma IL-4 (Pre-Ex: 0.24 ± 0.03 pg/ml, Post-Ex: 0.33 ± 0.04 pg/ml; ES= 0.51, p= 0.0330, Figure 4B) was significantly elevated compared to the baseline value. The peripheral IL-13 level (Pre-Ex: 10.84 ± 0.66 pg/ml, Post-Ex: 11.90 ± 0.95 pg/ml; ES= 0.27, p= 0.0632, Figure 4C) was borderline significantly increased after the exercise regimen.

**Figure 4.**
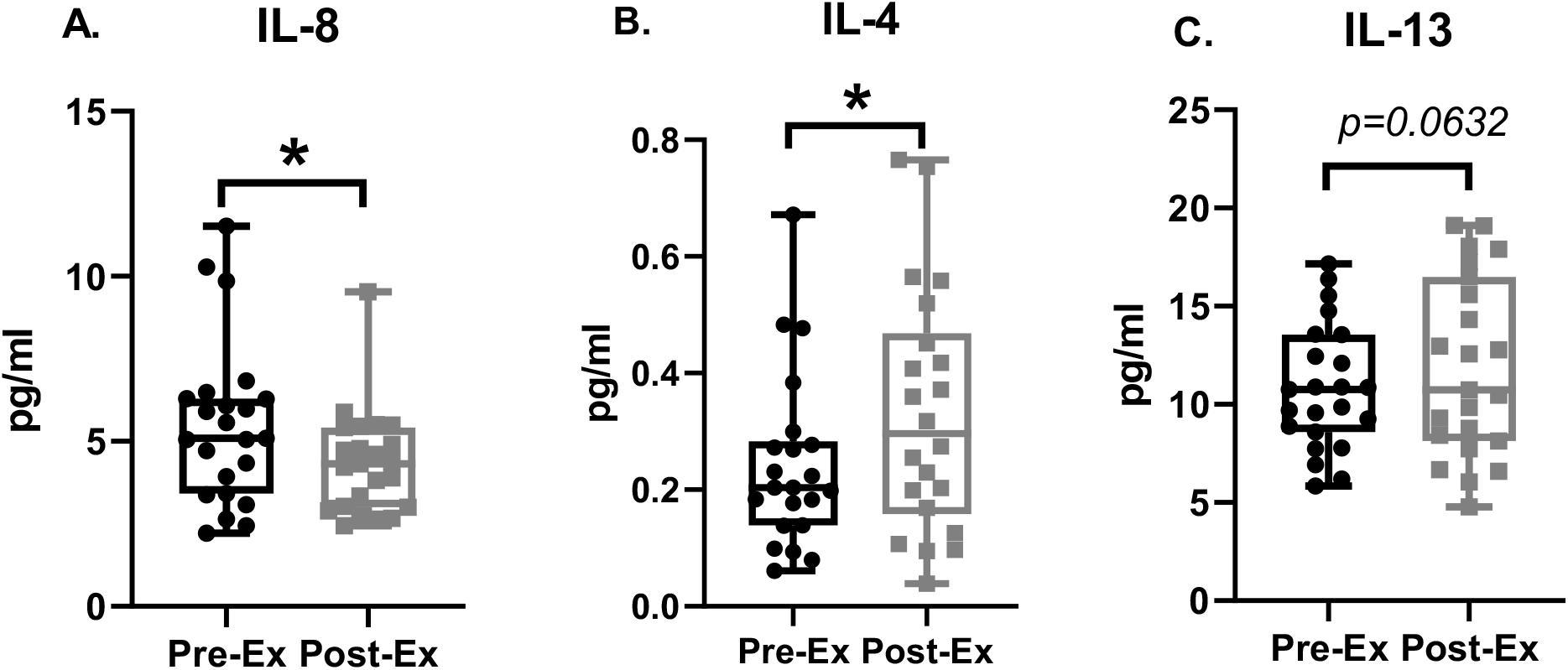
Comparison of baseline (pre-ex) and after 12-week exercise (post-ex) plasma cytokines level, interleukin-8 (IL-8, Fig. 4A), IL-4 (Fig. 4B) and IL-13 (Fig. 4C). N=24, data is mean ± standard error. * is set as statistically significant, p<0.05.

### BAI and sleep micro-architecture

No differences were seen in BAI measured through in-lab PSG at baseline and post-exercise timepoints (Pre-Ex: 0.87 ± 1.89 years, Post-Ex: 0.69 ± 1.83 years; p= 0.9026; Figure 5A). Additionally, we did not observe significant changes in sleep micro-architectural features, including delta band power during N3 sleep (Pre-Ex: 1.31 ± 0.09 dB, Post-Ex: 1.31 ± 0.09 dB; p= 0.9734; Figure 5B) and alpha band power in the awake state (Pre-Ex: 0.27 ± 0.09 dB, Post-Ex: 0.23 ± 0.04 dB; p= 0.5238; Figure 5C). Likewise, no change was observed in spindle density (Pre-Ex: 2.37 ± 0.19, Post-Ex: 2.35 ± 0.18; p= 0.8420; Figure 5D).

**Figure 5.**
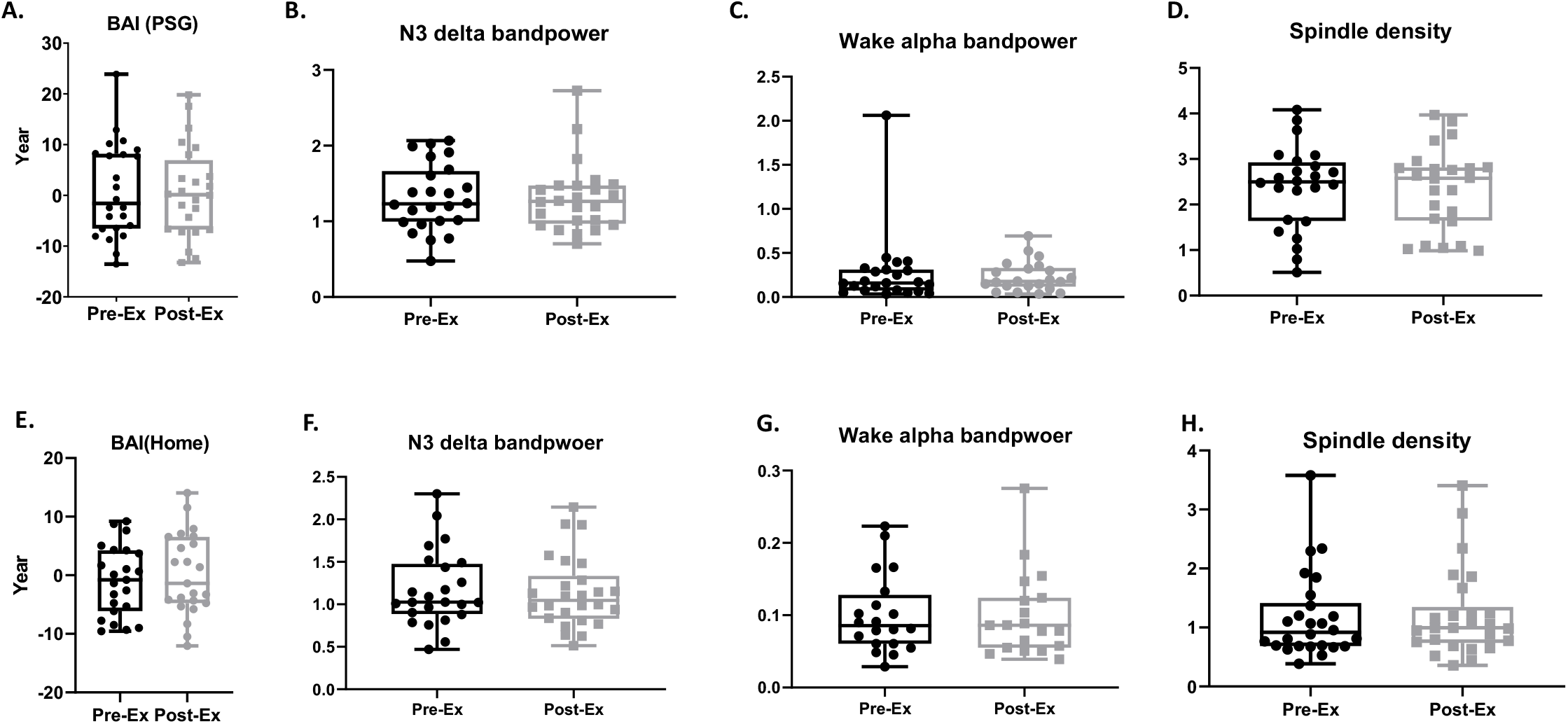
Comparison of baseline (pre-ex) and after 12-weeks of exercise (post-ex) brain age index and sleeping micro-architectures. Polysomnography (PSG) data, N=24 (Fig. 5A-5D) and home device data, N=26, (Fig. 5E-5H). Data is mean ± standard error.

After artifact detection and removal, 64% of home sleep headband data exhibited sufficient quality (artifact ratio < 0.5) to compute BAIs. As with in-lab PSG findings, no significant differences were seen in BAI measured by home sleep EEG when comparing baseline to post-exercise data: (Pre-Ex: -0.95 ± 1.24 years, Post-Ex: 0.13 ± 1.46 years; p= 0.3652; Figure 5E). Similarly, we found no significant changes in delta band power during N3 sleep (Pre-Ex: 1.17 ± 0.09 dB, Post-Ex: 1.13 ± 0.08 dB; p=0.6447; Figure 5F), alpha band power in the awake state (Pre-Ex: 0.1 ± 0.05 dB, Post-Ex: 0.09 ± 0.06 dB; p=0.6849; Figure. 5G), or in spindle density (Pre-Ex: 1.17 ± 0.14, Post-Ex: 1.21 ± 0.15; p=0.4585; Figure 5H).

### Sleep macro-architecture

We analyzed conventional metrics of sleep quality based on sleep macro-architecture from in-lab PSG and home sleep device data, to determine whether the 12-week exercise regimen was able to improve sleep quality. We found no significant differences in sleep stage percentages in the pre- vs post-exercise in-lab PSG data (Figure 6A, B) for the wake (Pre-Ex: 16.63 ± 2.46%, Post-Ex: 17.83 ± 2.12%; p=0.6498), REM (Pre-Ex: 16.63 ± 2.46%, Post-Ex: 21.31 ± 2.80%; p=0.8107), and NREM states (Pre-Ex: 62.49 ± 3.39%, Post-Ex: 60.86 ± 3.06%; p=0.5982). Similarly, we found no significant differences in stage percentages for sleep measured with home EEG (Figure. 6A and B) for wake (Pre-Ex: 21.27 ± 2.3%, Post-Ex: 21.18 ± 2.57%; p=0.9659), REM (Pre-Ex: 19.95 ± 1.45%, Post-Ex: 19.94 ± 1.14%; p= 0.9932), and NREM (Pre-Ex: 57.99 ± 2.15%, Post-Ex: 58.53 ± 2.15%; p= 0.7493).

**Figure 6.**
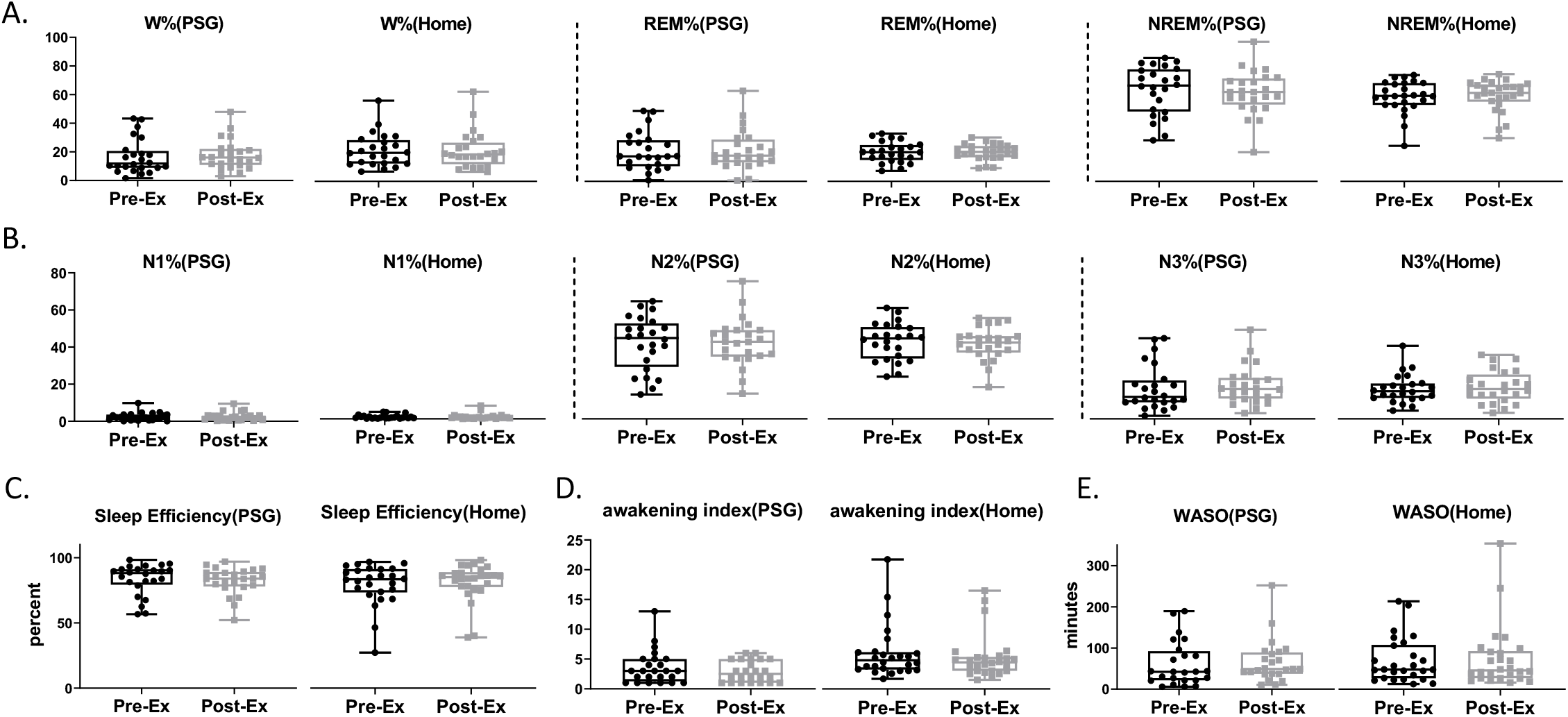
Trend of 12-week (x-axis starting from first day to last day of using home device) brain age index (BAI, Fig. 6A), non-rapid eye movement stage 3 (N3 percentage, Fig. 6B) and sleeping heart rate (HR beats per minute, Fig. 6C), collected from home devices. P-value was analyzed by robust linear model (RLM, green line). Ordinary least squares (OLS, red line). 95% confidence interval (shaded area). N=26.

We also found no significant exercise effects on sleep quality metrics derived from sleep macro-architecture measured with in-lab PSG (Figure 6C), including sleep efficiency (Pre-Ex: 83.37 ± 2.46%, Post-Ex: 82.17 ± 2.12%; p= 0.6498), awakening index (Pre-Ex: 3.58 ± 0.58, Post-Ex: 2.91 ± 0.36; p= 0.2592), and wake after sleep onset (WASO) (Pre-Ex: 61.52 ± 11.11 mins, Post-Ex: 66.58 ± 10.77 mins; p= 0.7250). Similarly, no exercise effects were found in home sleep data (Figure 6C) for sleep efficiency (Pre-Ex: 76.8 ± 2.99%, Post-Ex: 78.27 ± 2.72%; p= 0.3697), Awakening index (Pre-Ex: 5.96 ± 0.86, Post-Ex: 5.26 ± 0.72; p= 0.2160), and WASO (Pre-Ex: 67.27 ± 10.39 mins, Post-Ex: 71.5 ± 14.21 mins; p= 0.5903).

### Trends in BAI, sleep micro-architecture, and sleeping heart rate over 12 weeks of exercise

Based on the sleep EEG and heart rate data collected from the home wearable devices, we found that BAI was stable (p=0.9446, Figure 7A) throughout the exercise program. The percentage of N3 sleep showed a significant upward trend (p=0.0083, Figure 7B). Sleeping heart rate showed a significant downward trend (p=0.0251, Figure 7C).

**Figure 7.**
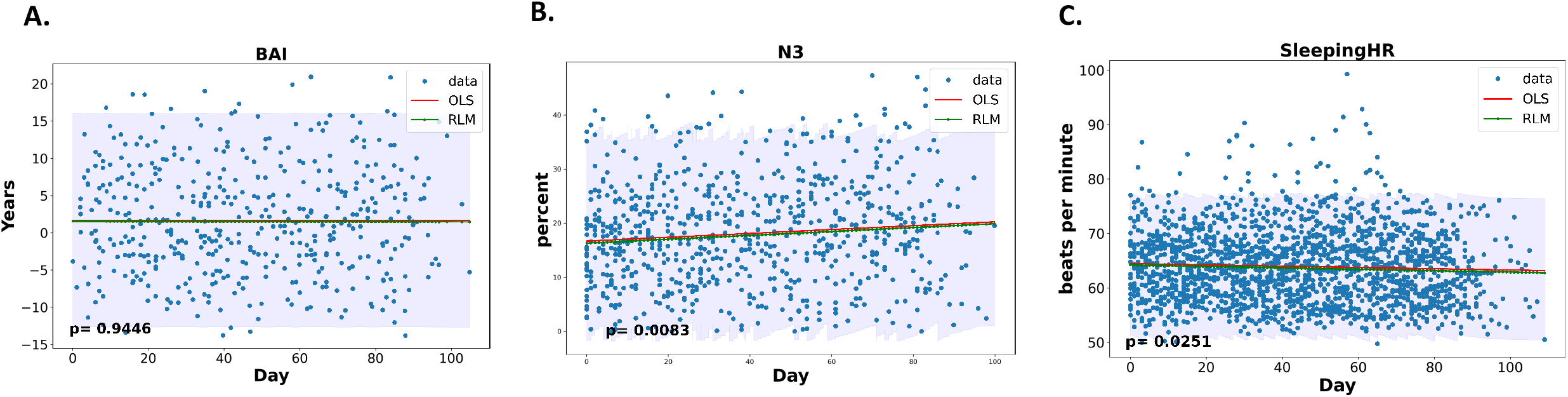
Comparison of baseline (pre-ex) and after 12-weeks of exercise (post-ex) conventional metrics of sleep quality. Percents of sleep stage: wake, rapid eye movement, non-rapid eye movement (W, REM and NREM, Fig. 7A), N1, N2, and N3 (Fig. 7B). Sleep fragmentation data: sleep efficiency (Fig. 7C), awakening index (Fig. 7D) and wake after sleep onset (WASO, Fig. 7E). Data is mean ± standard error. PSG data (N=24; Home device data (N=24).

### Associations of the study outcomes

We further analyzed the relations between the study outcomes by *Pearson’s* correlation (Table 2, Figure S2). The improvement of VO_2_max was positively (p=0.0050) related with the enhancement of cognition performance while not related with the change of BAI (p=0.8914)). The excess of BAI was related with less NREM (p=0.0035) sleep and more fragmented sleep as measured by wake stage percent (p=0.0099), sleep efficiency (p=0.0099) and WASO (0.0252). In addition, the increment of BAI was associated with lower level of plasma IL-4 (p=0.0240) and IL-13 (p=0.0023). The improvement of cognitive executive function measured by dimension card change sort was positively related with increased N2 (p=0.0150) and NREM (0.0370) sleep percentages.

**Table 2.** The associations between the change of aerobic fitness, cognition functions, sleep micro- and macro-architecture and plasma cytokines. Pearson’s correlation to verify the associations between the change of aerobic fitness, cognition functions, sleep micro- and macro-architecture and plasma cytokines. Maximal oxygen consumption: VO2max. SHR: sleeping heart rate. FLD: cognition fluid. DCCS: dimension change card sort. FICA: flanker Inhibitory control and attention. LSWM: list sort working memory. PCPS: pattern comparison processing speed. ORR: oral reading recognition. PVT: picture vocabulary test. BAI: brain age index. WASO: wake after sleep onset. NREM(N): Non-rapid eye movement. IL-4, 8, and 13: Interleukin-4, 8 and 13. N=24.

|  | **Features** | **Pearson's *r*** | ***p* value** |
| --- | --- | --- | --- |
| **VO<sub>2</sub>max vs.** | SHR | -0.4700 | **0.0160** |
|  | FLD | 0.6500 | **0.0003** |
|  | Cognition Total | 0.5300 | **0.0050** |
|  | DCCS | 0.6200 | **0.0007** |
|  | FICA | 0.4900 | **0.0115** |
|  | LSWM | 0.7000 | **<0.0001** |
|  | PCPS | 0.4000 | **0.0441** |
|  | BAI | 0.03 | 0.8914 |
| **BAI vs.** | sleep efficiency | -0.5300 | **0.0099** |
|  | WASO | 0.4700 | **0.0252** |
|  | Wake | 0.5300 | **0.0099** |
|  | N2 | -0.4400 | **0.0368** |
|  | N3 | -0.4200 | **0.0440** |
|  | NREM | -0.5800 | **0.0035** |
|  | IL-13 | -0.6000 | **0.0023** |
|  | IL-4 | -0.4700 | **0.0240** |
| **DCCS vs.** | N2 | 0.5010 | **0.0150** |
|  | NREM | 0.4370 | **0.0370** |
| **IL-13 vs.** | PVT | -0.4700 | **0.0246** |
|  | sleep efficiency | 0.4600 | **0.0262** |
|  | Wake | -0.4600 | **0.0262** |
|  | N3 | 0.5500 | **0.0062** |
|  | NREM | 0.5400 | **0.0081** |
| **IL-4 vs.** | PCPS | 0.5100 | **0.0134** |
|  | ORR | 0.4300 | **0.0412** |
|  | delta band power in N3 | 0.4200 | **0.0435** |
| **IL-8 vs.** | awakening index | 0.4400 | **0.0341** |

Additionally, we found higher level of IL-13 and IL-4 and lower level of IL-8 were significantly related with better sleep quality as measured by sleep efficiency (p=0.0262), N3 percentage (p=0.0062), NREM percentage (p=0.0081), awakening index (p=0.0341) and delta band power during deep sleep (p=0.0435). Higher level of IL-4 was also significantly associated with the improvement of cognition performance measured by processing speed and language ability.

Results of the linear mixed effects modeling to analyze the relations between BAI, VO_2_max, heart rate, cognitions and plasma cytokines, adjusted for age and sex, as shown in Table 3. The results were computed from the whole dataset including the five withdrawn participants who only completed baseline assessments. Resting heart rate was negatively associated with VO_2_max (p=0.03). Moreover, BAI predicted from PSG data was positively related with plasma IL-8 (p=0.035), and inversely related with fluid intelligence (p=0.009), processing speed (p=0.002), IL-4 (p=0.009), IL-13 (p<0.0001) and potentially memory performance (p=0.103). Additionally, we found a trend between PSG-derived BAI and resting heart rate, a predictor of VO_2_max (p=0.087). However, BAI calculated from PSG data was not correlated directly with VO_2_max (p=0.564). We did not observe any significant relations between BAI calculated from home EEG data and VO_2_max, cognitive performance measures, heart rate, or cytokine levels.

**Table 3.** Mixed Effects Model. Mixed linear regression model to calculate the associations of all pre- and post-exercise data. Var.: variable. VO_2_max: maximal oxygen consumption. BAI: Brain age index. HR: Heart rate. FLD: Cognition fluid. PCPS: pattern comparison processing speed. LSWM: list sort working memory. IL-4, 8 and 13: Interleukin-4, 8 and 13. N=31.

| **Dependent var.** | **Independent var.** | ***p* value** |
| --- | --- | --- |
| VO<sub>2</sub>max | BAI | 0.564 |
|  | Resting HR | **0.03** |
| BAI | VO<sub>2</sub>max | 0.868 |
|  | Resting HR | 0.087 |
| FLD | BAI | **0.009** |
| PCPS |  | **0.002** |
| LSWM |  | **0.103** |
| IL-4 |  | **0.009** |
| IL-8 |  | **0.035** |
| IL-13 |  | **<0.0001** |

## Discussion

This interventional clinical trial evaluated the efficacy of a 12-week moderate-intensity exercise program to improve sleep and brain health in sedentary older adults without sleep complaints. Compared to previous studies that rely on subjective sleep reports or coarse measures of sleep, our study collected abundant objective physiological data in both in-lab and home settings. Our results suggest that 1) N3 sleep percentage, plasma cytokines, cognitive performance and physical fitness were improved in sedentary older adults after a 12-week moderate-intensity aerobic exercise regimen; 2) BAI, as currently computed, was not sensitive enough at our sample size to detect neurophysiologic correlates of these improvements in cognition for this type of population after this level of improved physical fitness; 3) the improvement of cognitive performance was associated with improvements in aerobic fitness, N2 and NREM sleep percentages and higher circulating IL-4/13, although not with BAI; and 4) lower BAI was associated with less fragmented sleep and higher levels of neuroprotective cytokines (IL-4/13). These findings demonstrate that moderate intensity aerobic exercise over a relatively short period may improve brain health and sleep quality in previously sedentary older adults.

### Improvements in cognitive performance

We found that 12-week, 150 minutes per week of moderate-intensity exercise produced measurable improvements in fluid intelligence, crystallized intelligence, and total cognitive performance. Specifically, processing speed and oral reading recognition skill were moderately improved, and improvements in working memory were marginally significant. Our results agree with previous reports showing moderate improvements in processing speed (∼7%) following aerobic exercise in older adults [46, 47]. Moreover, our data follows the agreement of a recent study that 12-week strength training improves fluid cognition in older adult [48]. In contrast to processing speed, previous studies show mixed results regarding working memory improvement following aerobic exercise [46, 49, 50], likely due to different sample demographics and assessment methods for working memory. However, one meta-analysis found that healthy participants exhibit improvements in working memory following exercise, in line with our finding [50]. Further, to determine whether the improvement of cognition performance is related to learning or practice effect, we compared the effect size of our cognition performance with a validation study results [34]. We found that the current 12-week exercise program produced higher effect size of cognition scores compared to the 7 to 21 days test-retest effect. Additionally, the 12 week interval is long enough to minimize the test-retest practice effect of cognition function [51]. Therefore, we rationally advocate the improvements of cognition performance is the result of 12 weeks aerobic exercise regimen.

There may be multiple mechanisms by which a moderate-intensity exercise regimen improves cognitive performance, including : 1) improved cerebrovascular function [52, 53]; 2) increase in hippocampal volume [54] and reduction in the burden of white matter lesions [55], 3) regulation of peripheral cytokines and neurotrophic factors such as, Brain-Derived Neurotrophic Factor (BDNF), [56, 57], IL-6, cathepsin B, irisin [58] and IGF-1 [59]. Here, we found that several plasma cytokines, IL-4, IL-8, and IL-13 were altered after the exercise regimen. Importantly, we further observed that the improvement of processing speed and oral reading recognition skill were related with the increment of IL-4. Our results reinforce the key functions of anti-inflammatory cytokines IL-4/13 which have been shown to exert neuroprotective effects on activated microglia to protect neurons from injury and hippocampus volume loss [60-62]. Moreover, higher level of IL-4 in both animal and human trials is associated with better cognitive performance [63, 64] while increased circulating proinflammatory IL-8 is associated with lower memory and processing speed [65]. The observation of reduced circulating IL-8 over the 12-week exercise program is consistent with the finding of a different exercise regimen which showed a rapid decrease of IL-8 30-mins after exercise [66]. Additionally, IL-8 is strongly associated with oxidative stress [67, 68] which is associated with impaired cognition function [69]. Thus, our data suggest that one mechanism for the observed therapeutic effects of exercise on brain cognitive health may be improvements in circulating plasma cytokines. It might also be of interest in the future to measure the effects of physical exercise on biological aging using an DNA methylation epigenetic clock in parallel with the EEG based BAI [70]. The findings of our current study warrant future research to further characterize the mechanisms by which exercise improves cognitive functions.

### Sleep and sleep EEG based brain age index

When evaluating brain health through measures of BAI and other sleep metrics, we did not observe changes between baseline and post-exercise training evaluations. Similar to previous work, the effects of exercise training on sleep macro- or micro-architectures remain uncertain, especially in people without sleep complaints [71]. For example, acute exercise training has been reported in some studies to increase delta band power [72, 73] and slow wave sleep duration [74]. However, another randomized study concluded that a 12-month moderate-intensity exercise program improved several objective measures of sleep but did not change measures of slow wave sleep [9]. Multiple factors (e.g. exercise intensity, duration, the time before sleep when exercise was completed, night-to-night variability, and the individual differences in participants) could potentially explain these inconsistencies. In the present study, a trend across the 12-week intervention period showed an increase of 3.59% in percentage of N3 sleep, which provides some evidence that 12-week moderate-intensity exercise may improve sleep quality. We further found that younger brain age (lower BAI) was associated with more N2 and N3 percentages of sleep and less fragmented sleep, supporting the validity BAI as an indicator of brain health, despite the lack of measurable changes induced by exercise over the course of the 12-week study. Additionally, we observed that higher plasma IL-13 is significantly associated with increased sleep efficiency and increased slow wave sleep as a percentage of total sleep, while higher IL-8 level was related with more fragmented sleep. Our findings are in agreement with previous work that circulating IL-8 is significantly increased in sleep disordered patients compared to healthy controls [65]. Moreover, the higher levels of the neuroprotective cytokines IL-4/13 were significantly with younger brain age, which is the first time an association has been shown between circulating biomarkers and BAI. Furthermore, we discovered that circulating IL-4 is positively correlated with slow wave sleep (SWS) power during N3 sleep. Recently, there has been increasing interest in augmenting SWS to improve cognition, such as through transcranial electrical stimulation [75, 76] and controlled acoustic stimuli [77]. SWS and increased slow wave power during sleep is associated with several desirable physiological effects, including enhanced glymphatic flow [78, 79], stable breathing and vagal dominance of heart rate variability [80]. Along with the relationship between sleep microarchitecture and brain cognition health, we observed that better brain executive function is associated with more N2 and NREM sleep macro-architecture, which is similar to previous finding that improved NREM sleep contributes to execution functioning [81]. As aforementioned, the higher levels of IL4/13 are associated with improved cognitive health.

It is also noteworthy that this study was completed during the COVID-19 pandemic. This global pandemic is strongly associated with widespread heightening of anxiety and depression levels [82, 83], significant changes in lifestyle (increase in daily sedentary time and nap time) [84], and increased disturbances in sleep patterns and quality [85, 86]. Therefore, the impact of COVID-19 on older adults, who are already at high risk of critical illness and morbidity, may be non-negligible and may have counteracted some benefits of the exercise program. In the present study, the changes in sleeping heart rate and improvements in exercise performance were modest. It is quite possible that a more vigorous and prolonged program may show changes not evident in our study. Selecting particularly physically deconditioned, not just sedentary, individuals may also show benefits not demonstrable in our population. Individual differences in brain macro- and micro-architecture during sleep may also be important, with exercise providing greater benefit in those with reduced baseline slow wave power.

### Improvements in aerobic fitness

As hypothesized, the 12-week moderate intensity aerobic exercise program increased VO_2_max and decreased resting and sleeping heart rate. Moreover, a 12-week trend analysis suggested sleeping heart rate decreased by approximately 1.3 beats per minute per 100 days. Combined with the SWS trend described above, the longitudinal home data illustrates how sleep quality and heart function responded to exercise over time. We found that improvements in VO_2_max were associated with improvements in cognitive performance and reduction of sleeping heart rate, but not with change in BAI and sleep macro- and micro-architecture. Additionally, we didn’t find any associations between the improvement of VO_2_max and the change of expression of plasma cytokines either. Our data generally aligns with previous findings that describe increases in physical fitness and brain health following aerobic exercise [87-89] and a link between elevation of exercise capacity with improvements in brain cognitive health, suggesting that the current intensity and frequency of workouts promote brain health.

Finally, our mixed effects model result suggested a lack of association between BAI and VO_2_max, but significantly correlations between BAI, fluid intelligence processing speed, IL4, IL-13 and IL-8. Processing speed performance changes over the lifespan, peaking during early adulthood and decreasing from there on [90, 91]. Importantly, processing speed is among the most sensitive cognitive processes influenced by neurological illness [92, 93] and is able to differentiate healthy populations from dementia populations [94, 95]. Our previous work demonstrated that BAI is sensitive to the severity of dementia [30]. Additionally, IL-4 and IL-13 are the critical anti-inflammatory cytokines to regulate sleep [96] and cognition performance [97]. Collectively, our study shows promise in this line of research, and further work focusing on long-term effects of exercise and lifestyle modifications may support or improve on BAI to create a noninvasive physiological biomarker that reliably measures brain health and its response to interventions.

### Limitations

The lack of change pre- and post-exercise training seen for BAI and various sleep metrics might be due to suboptimal exercise regimen in both intensity and duration, ceiling-floor effects due to our relatively healthy sample [98], small sample size, and the night-to-night variability of sleep measures including BAI [99]. As described in the Methods, we initially targeted a sample size of N = 35 but had to stop enrollment due to unanticipated challenges of the COVID-19 pandemic, resulting in a loss of power. This is one major limitation which may contribute to the negative findings regarding BAI. Another major limitation is the limited data quality of home sleep EEG devices. We employed home sleep headbands to record multiple nights of sleep EEG to account for night-to-night variability of BAI calculations. However, after artifact detection, 64% of home sleep EEG data had an artifact ratio <0.5 and only 44% had an artifact ratio <0.3. Though, despite collecting abundant longitudinal home sleep EEG data, only a portion of this was suitable for analysis, limiting our ability to average over multiple close-together nights to detect more subtle changes in BAI. Another limitation is most participants in the study are white college educated female. These sociodemographic factors should be aware of in future work.

## Conclusion

These data provide evidence that a 12-week, 150 minutes weekly moderate-intensity, aerobic exercise regimen improves cognitive performance and slow wave sleep duration in association with improved aerobic fitness and higher circulating neuroprotective cytokines among older previously sedentary adults.

## Data Availability

The data in the manuscript will be available as requested

## Author contributions

Conceptualization: AO, HS, LP, CZ, JR, RET, MBW

Methodology: AO, HS, NA, RAT, DL, PF, WG, JJ, LG, CZ, RET, MBW

Investigation: MB, JR, RET, MBW

Project administration: NA, RAT

Writing – original draft: AO, CZ, NA, RAT, HS, CZ, RT, JR, RET, MBW

Writing – review & editing: AO, CZ, NA, RAT, HS, DL, JJ, WG, JS, MB, CW, ZC, GF, RT, JR, RET, MBW

